# A Systematic Review of Measles Virus Transmissibility in the Air to Guide Exposure Periods for Contact Tracing in Public Spaces

**DOI:** 10.1101/2025.03.26.25324620

**Authors:** Haley F. Farrie, Sarah Gillani, Fatima Nkempu Ameaka, Elizabeth Campbell, Eric S. Toner, Sanjana Ravi, Sutyajeet Soneja, Caitlin M. Rivers

## Abstract

**Objectives:** To systematically review epidemiological evidence to determine how long the measles virus remains transmissible after an infectious case leaves a public space to address inconsistencies in measles contact tracing exposure window guidelines.

**Methods:** A systematic literature review following PRISMA guidelines was conducted using PubMed, EMBASE, Web of Science, and SCOPUS databases for publications from January 1988 to July 2024. Additional sources were identified through reference list reviews and Google Scholar searches. Studies examining how long the measles virus survives in the air or remains transmissible after an infectious case leaves a public space were included, while non-evidence-based recommendations and mathematical models were excluded.

**Results:** Initial database searches identified 1,054 studies, with none meeting initial inclusion criteria after screening. Supplemental searches identified five relevant articles (1964-1987). Two experimental studies demonstrated measles virus survival between 30 and 120 minutes, with increasing survival time for lower humidity levels.

Experimental studies showed survival at two hours in low humidity (12-15%), up to 60 minutes in moderate humidity (36-37%), and 30 minutes in high humidity (60-70%) at 20°C. Three publications reviewed how long measles virus was transmissible in real-world settings, ranging from 60 to 120 minutes.

**Conclusions:** Limited evidence exists to guide the precise determination of the duration of measles transmissibility. Current health department guidelines rely on limited research from 1964 to 1987. Additional studies are needed to understand how long the virus is transmissible in real-world settings, particularly given the implications for contact tracing efficiency and resource allocation during outbreak responses.

## Introduction

The United States eliminated measles in 2000^1^ due to high vaccination rates and effective public health measures; however, the U.S. has experienced several outbreaks since then, typically after introduction from international travelers.^2^ These situations are managed by health department teams, which act quickly to identify people exposed to the case, provide post-exposure prophylaxis (PEP) to individuals without immunity where possible, and quarantine contacts at risk of developing the infection. These responses are resource-intensive, require significant staff time and funding, and can detract from routine health department operations.^3^

Contact tracing for measles presents unique challenges. Like influenza^4^ and SARS-CoV-2^5^, measles spreads through airborne and droplet transmission.^6^ However, measles also remains infectious in the air for an extended time, even after the infected person is no longer present.^7^ According to the current U.S. Centers for Disease Control and Prevention (CDC) measles exposure guidelines, if a person experiences either of the following without personal protective equipment for any amount of time, they are considered exposed: 1) occupying the same air space at the same time as someone with measles or 2) entering a room within 2 hours after a person with measles left.^7^ These exposure guidelines are based on two experimental studies of how long the measles virus remains in the air^8,9^, three publications reviewing the evidence of real-world outbreaks (1985-1987)^10–12^, and a literature review.^13^

The Johns Hopkins Center for Outbreak Response Innovation conducted eleven key informant interviews with U.S. state and local health departments to understand the operational challenges related to contact tracing and outbreak response. Interviews took place between March and October of 2024. These interviews revealed variations in the measles exposure guidelines for contact tracing.^14^ While most health departments utilize the two-hour exposure window recommended by the CDC, two relied on a more lenient one-hour exposure window.^15^ The exposure window directly impacts contact tracing procedures. Reducing the exposure window from two hours to one hour can substantially reduce the number of people considered exposed and thus reduce the number of contacts requiring notification/quarantine. A shorter exposure window would likely considerably reduce staff hours and costs required to conduct these activities.^16^ Moreover, accurately identifying the exposure window facilitates efficient contact tracing within the 72-hour window when PEP remains effective.^17^ Accurately identifying this timeframe is also crucial for ensuring that staff time and funding are appropriately managed and that routine health department work is not disrupted unnecessarily. To address this inconsistency and ensure that exposure guidelines are based on the strongest data available, we conducted a systematic review of the evidence for how long the measles virus remains airborne and capable of infection after an infectious case has left an area.

## Methods

This systematic literature review follows the Preferred Reporting Items for Systematic Reviews and Meta-Analyses (PRISMA) guidelines.^18^ This study aimed to examine the duration of measles virus infectivity in airborne environments to guide epidemiologists in determining exposure periods for contact tracing in public spaces after the presence of an infectious case.

### Literature Search Strategy

The literature search was conducted in Pubmed, EMBASE, Web of Science, and SCOPUS for articles published between January 1988 and July 2024. This starting year was chosen as it marks the commercialization of the taq polymerase enzyme critical for the widespread utilization of polymerase chain reaction (PCR) technology for accurate measles diagnosis.^19^ A search query was developed with expert input utilizing mesh database terms. The following search query was used: (“measles” OR “rubeola”) AND (“air microbiology” OR “respiratory aerosols and droplets” OR “airborne” OR “exposure period” OR “survival time” OR “environmental stability” OR “aerosol*” OR “droplet*” OR “air”) AND (“disease transmission, infectious” OR “transmission” OR “airborne transmission” OR “survival” OR “mortality”). All results were managed through Covidence^20^, a systematic review processing software, for de-duplication, title and abstract screening, full-text review, and extraction.

### Study Screening and Eligibility

After de-duplication, two independent reviewers screened titles and abstracts to assess eligibility for full-text review. Any disagreements were resolved by a third independent reviewer, who made a final determination.

Studies were included if they examined the duration of measles infectivity in the air after an infected person has left a space, specifically in public places (e.g., hospitals, airplanes, grocery stores). Studies that occurred indoors and outdoors were both eligible for inclusion. Eligible studies included peer-reviewed articles and grey literature from public health authorities that address contact tracing in relation to exposure periods and review the relevant scientific evidence. Both human and animal studies, as well as laboratory-based investigations on measles infectivity in the air, were considered.

Exclusion criteria included recommendations and studies that were not evidence-based (including mathematical models), unrelated to airborne transmission of measles in public environments, and publications not in English. Reviews, editorials, opinion pieces, and duplicate studies or studies with overlapping data sets were also excluded. If an outbreak or airborne transmission event was discussed in both peer-reviewed articles and grey literature, for example, journal publications, only the peer-reviewed articles were included to reduce duplicative reporting of results.

The manuscripts that met inclusion criteria during the title and abstract screening were advanced to a full text review. During this phase, two reviewers again independently assessed each manuscript for inclusion. Items that received discordant votes were each discussed during a live meeting attended by both primary reviewers and a third reviewer. In this meeting, each reviewer presented their rationale, and the third reviewer made the final determination after considering all viewpoints. Any manuscripts that passed the full-text review were subsequently included in the data extraction and analysis phase.

### Supplemental Search Strategies

Due to the dearth of results from the systematic literature search, additional search strategies were conducted to ensure comprehensive coverage of the evidence. These three additional methods included 1) review of references from literature reviews identified during the Covidence search, 2) a Google Scholar search utilizing the same search query as was implemented in the study databases, and 3) a review of all literature used by the U.S. CDC to inform the established measles exposure guidelines. These supplemental search strategies followed the same inclusion and exclusion criteria used during the systematic search, except that there was no limitation applied to the date of publication.

Literature reviews, and therefore their content, identified during the systematic search were originally excluded because they did not meet the original search criteria for primary data collection. However, any literature review references that specified primary data collection of measles exposure data with timeframes were included as part of the supplemental search; this is referred to later in this write-up as the literature review snowball sampling. Although the literature reviews themselves are still not included in the results, the primary research articles themselves are included. The second supplemental search strategy was the Google Scholar search, which was conducted using key phrases searched in combination and independently: “Measles Exposure,” “Measles Outbreak,” “Measles Case Study,” “Measles Case”, “Measles Evidence,” “Measles Airborne,” and “Measles Transmission.” The third strategy involved reviewing the evidence published on the U.S. CDC website.^7^ To aid the supplemental search, the findings of this systematic review were also shared with the state and local health department partners identified through previous interviews, including those that followed both one- and two-hour exposure windows, and no additional articles were added.^14^

### Data Extraction

Data from studies identified in the supplemental search were extracted into a summary table (Table 1) to outline the findings, authors, and any key factors related to the findings. This table includes the setting, whether it was an experimental laboratory setting, a provider’s office, or a hospital pharmacy, the environmental conditions (humidity and room temperature if reported), and the infectious exposure window.

**Table 1.** Summary of all publications utilizing primary data that could be used to inform exposure windows for measles contact tracing after an infectious case is no longer present in indoor or outdoor space.

| Setting | Environmental Conditions | Infectious Exposure Window | Citation |
| --- | --- | --- | --- |
| Laboratory Experiment | Humidity: 12-15%<br>Temperature: 20°C | 120 min | De Jong et al. 1964 <sup>a</sup> |
|  | Humidity: 68-70%<br>Temperature: 20°C | <30 min |  |
|  | Humidity: 36-37%<br>Temperature: 20°C | >60 min | De Jong et al. 1965 <sup>b</sup> |
|  | Humidity: 60-64%<br>Temperature: 20°C | 30 min |  |
| Pediatric Outpatient Office | Humidity: 22%<br>Temperature: 20°C | 60-75 min | Remington et al. 1985 <sup>a</sup> |
|  | Humidity: not reported<br>Temperature: not reported | 1hr | Bloch et al. 1985 <sup>a</sup> |
| Hospital Pharmacy | Humidity: not reported<br>Temperature: not reported | 29-120 min | Sienko et al. 1987 <sup>a</sup> |

<sup>a</sup> Utilized to inform CDC measles contact tracing guidelines.
<sup>b</sup> Not utilized to inform CDC measles contact tracing guidelines.

### Quality Assessment

A standardized quality assessment tool^21^ was utilized to ensure the reliability and validity of the data identified through the systematic review. The mixed methods appraisal tool (MMAT) was used to allow for flexibility in the review of the study designs captured through the systematic search. The methodological quality criteria for quantitative non-randomized studies were used to best assess the clarity of the research questions, the correctness and quality of the data used to address the research questions, the representativeness of the target population, appropriate measurement of the outcome and exposure, completeness of outcome data, appropriate accounting of confounders in the design and analysis, and proper measurement of the exposure. The 7-question quality assessment was completed for every article that was included in the data extraction. Quality assessments were conducted blinded by two independent reviewers and any disagreements were discussed and resolved.

## Results

### Data Extraction and Review

Database searches initially identified 1,054 studies from four databases: Scopus (392), Web of Science (262), PubMed (200), and Embase (200), as shown in Figure 1 below. After removing 495 duplicates, 559 studies remained for the title and abstract screening. After the first round of screening, 463 studies were excluded based on criteria outlined in the methodology, leaving 96 studies for full-text review. However, none of these studies ultimately met the previously mentioned criteria for data extraction and analysis.

**Figure 1.**
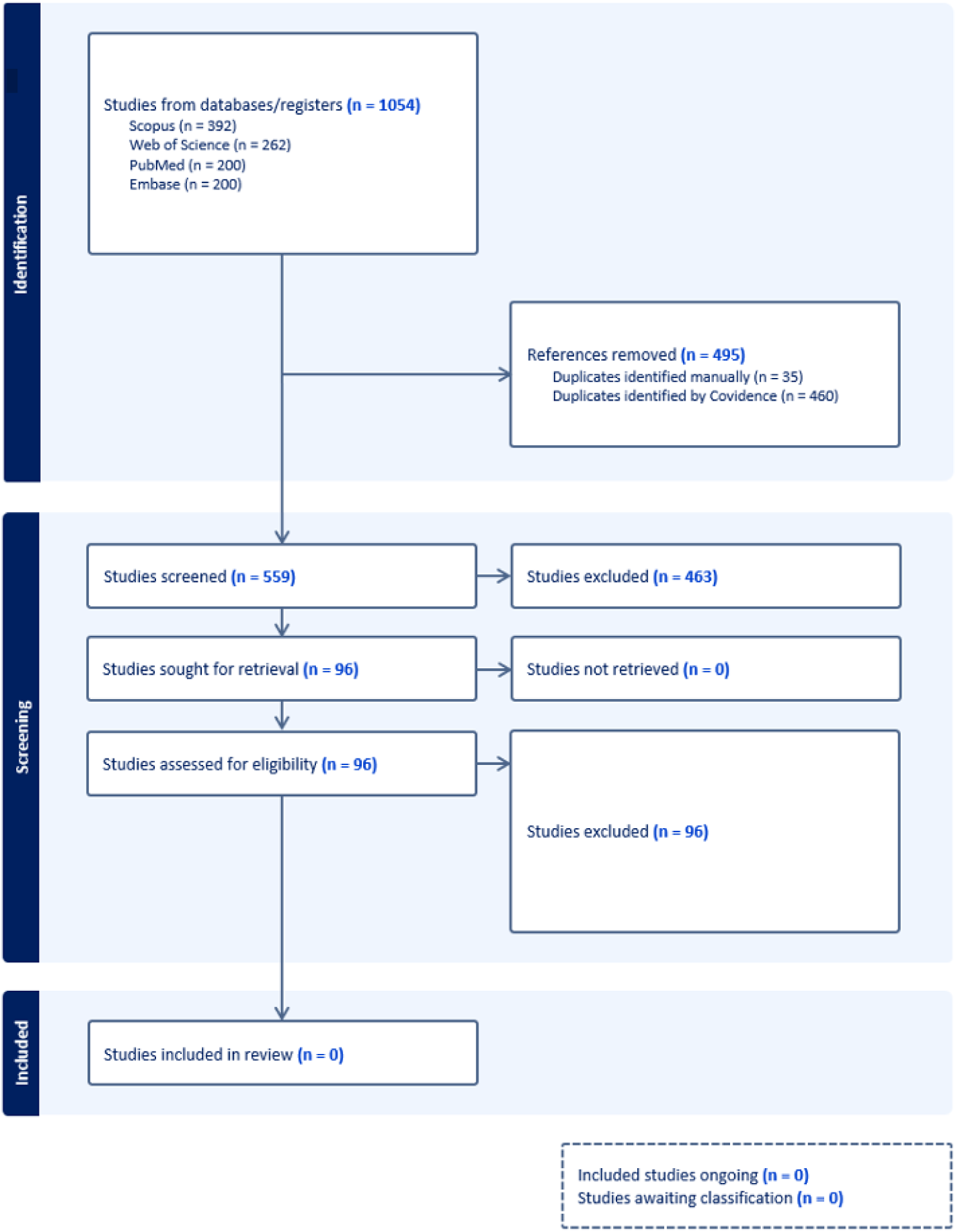
PRISMA Framework with the number of studies identified across each phase.

The supplemental search strategies employed identified only five articles containing evidence that could be used to determine how long measles virus remains infectious in the air after an infected person leaves a space. All five articles were published between 1965 and 1987, and two were published with the same lead author. None of the five studies contained primary data that could be used to inform measles contact tracing guidelines for outdoor spaces.

The first two publications assessing measles survival in the air were experimental studies published in 1964^8^ and 1965^9^. Measles virus survival in the air was used by the authors as a proxy for the window of time a person may be infected after an infected person leaves an indoor space. These studies used aerosolized measles virus in a room where air samples were taken at regular intervals and the surviving virus was estimated by titration. Both articles were published by the primary author J.G. de Jong one year apart. The first study, published in 1964, showed that measles virus survived two hours when disseminated in a room with low humidity (relative humidity 12-15%) but less than 30 minutes in a room with high humidity (relative humidity 68-70%).^8^ This study did not account for factors such as light or airflow, and temperature was held constant at 20°C (68°F). The second article, which was an extension of the first, considered additional factors that may affect the duration of measles infectivity in air, such as additional variations in humidity, slight variations in temperature, and the presence or absence of artificial light.^9^ These findings reaffirmed conclusions in the previous article and showed that measles virus survives for less time in the air with increasing levels of humidity when temperature is held constant at 20°C. In the 1965 study, measles was tested at 36-37% relative humidity (RH) and 60-64% RH, with the virus surviving for at least 60 minutes at the former RH and only 30 minutes at the latter RH. The presence or absence of artificial light was not found to have any effect on measles virus survival in the air when temperature and humidity were constant. Measles virus survival was slightly longer at 15°C than at 20°C, however, the influence of this on the survival time in the air was not reported.^9^

Twenty years later, three publications reported observational data that included evidence on the duration of measles virus transmissibility. The 1985 study by Remington et al. discusses an outbreak of measles in a pediatrician’s office. The article identified three children who developed measles after being exposed in a pediatric office 60 to 75 minutes after the index case had departed.^12^ According to the timeline provided by the authors, one of the secondary cases arrived at 1:30 PM (60 minutes after the case departed) and became infected; the next secondary case arrived a little after 1:30 PM (about 65 minutes after the case departed) and became infected; and the last secondary case arrived around 1:45 PM (about 75 minutes after the case departed) and became infected. Of the three children that became infected in the 60-75 minutes after the index case departed, two were under the age of 15 months and one was aged 15 months to 18 years. All three were unvaccinated. A total of 32 children were present within 90 minutes after the case departed, and only the three children discussed above became infected. An additional 28 children were present in the pediatric office 90 minutes or more after the infected case departed and none of these individuals developed a measles infection. Interestingly, the observed attack rates for unvaccinated children present with the index case and unvaccinated children that arrived less than 90 minutes after the index case are similar, around 25%, whereas the attack rate for unvaccinated children arriving 90 minutes or more after the index case is zero.

Remington et al. emphasized that factors such as coughing and inadequate fresh-air ventilation can prolong the duration of airborne infectivity, increasing the risk of transmission to susceptible individuals who enter the space after an infected person has left.^12^ The second observational publication by Bloch et al. describes a measles outbreak in a pediatric practice where the index patient, who was in the second day of rash, was able to transmit measles to other patients through airborne transmission, even after leaving the office.^11^ The article suggests that measles virus can remain infectious in the air for at least one hour after the index patient has left the area, providing evidence that measles can be transmitted through airborne spread even in the days after symptom onset, rather than just in the pre-rash period as previously believed. The specific duration and time windows are not reported as they had been in the Remington article; therefore, it is not clear how much greater than one hour the exposure window was. The final publication summarized how long measles virus remains transmissible after an infectious case leaves a space in a hospital setting.

During this outbreak, individuals who arrived 29 to 120 minutes after a case was present contracted measles. The minimum exposure window in this outbreak was observed when an 8-month-old was infected with measles after entering an emergency room 29 minutes after the index case was no longer present. The maximum exposure window observed was when a 24-year-old contracted measles while purchasing medications at the university hospital two hours after an infected internal medicine resident passed by the pharmacy seeking evaluation for her illness.^10^

A summary of the results is available in Table 1. Both the smallest and largest exposure windows were identified from laboratory studies. The smallest infectious exposure window reported was about 30 minutes, which was dependent on high humidity (68-70%) and room temperature (20°C) while the largest exposure window was about 120 minutes, which was dependent on low humidity (12-15%) at room temperature (20°C).^8^ Observational studies reported an exposure window within the range of that reported by the experimental studies, both within the general range of 60 to 90 minutes. No additional observational or experimental studies have met the criteria for assessing the exposure window since 1985.

### Quality Assessment

The 1964 experimental study by De Jong et al. provided the first published evidence that measles persists in the air for an extended period of time. The laboratory experiment presents clear research questions regarding measles virus survival in relation to relative humidity and seasonal morbidity patterns. While the experimental measurements of virus survival appear accurate, the study fails to account for important confounders such as temperature, airflow, and ventilation. The population data from England and Wales may limit its generalizability beyond the study’s geographic scope. Notably, the research does not address whether the surviving virus remains truly infectious in humans or examine specific exposure windows, which would be crucial for comprehensive understanding of transmission dynamics.

De Jong et al.’s 1965 follow-up experimental study expands on their previous work by including temperature and artificial light as variables alongside relative humidity. The measurements are appropriately designed with clear time points and exposures.

However, temperature data is inadequately reported; it was measured only at 15°C and 20°C with minimal detail and there was no visual representation in the article. While the study accounts for some potential confounders by individually exploring temperature and light effects, it still neglects airflow and ventilation factors. As with the previous study, the absence of human participants limits translation to real-world transmission scenarios.

The 1985 study by Remington et al. reported on a measles outbreak in a Muskegon, Michigan pediatrician’s office primarily affecting children and effectively documents the first evidence of measles virus transmissibility after a case leaves a space in a real-world setting. The research did this by tracking interactions between index cases and contacts under controlled environmental conditions (22% humidity, 20°C) and categorized exposure windows clearly (<90 minutes versus ≥90 minutes after index case departure), providing valuable data on transmission timing. Although demographic information for the 84 participants from Muskegon, MI is limited—noting only one infant Korean immigrant—the documentation of secondary cases is comprehensive. The study appropriately accounts for temperature, humidity, and airflow confounders, though additional information on exposure duration for each contact would have strengthened the findings. Additionally, the applicability of findings in light of recent modernization of ventilation systems is also in question.

Bloch et al.’s 1985 investigation into an outbreak in a pediatrician’s office in DeKalb County, Georgia also provides compelling evidence for measles virus survival in the air for at least one hour. Through meticulous documentation of exposure windows using appointment schedules, sign-in logs, and participant interviews, the researchers established that transmission occurred to a patient who entered a pediatric office one hour after the source patient had left. The study’s strength lies in its thorough airflow analysis, demonstrating that the virus spread through the ventilation system throughout the facility. While demographic information for the affected members of the DeKalb County population is limited, the researchers effectively ruled out fomite transmission and documented airflow dynamics, though humidity and temperature data were not captured.

The 1987 study by Sienko et al. clearly examines the duration of transmissible measles virus in a Michigan university medical setting. The research effectively documents exposure timeframes, enabling assessment of how long measles remains transmissible after a source patient departs. The place and time of exposure were well-recorded for all cases except one, which was still linked to an infected medical student. However, the study fails to account for potentially significant confounders including temperature, humidity, and ventilation characteristics. Demographic information is minimal, noting only that both children and adults were affected, which somewhat limits the generalizability of the findings.

The five studies collectively examined present valuable but limited evidence regarding measles virus transmission and survival, with methodological constraints that affect their applicability to contemporary settings. The De Jong et al. studies (1964, 1965) provide important foundational laboratory data but fail to address human infectiousness or account for all relevant environmental variables. The observational investigations from the 1980s (Remington, Bloch, and Sienko) offer useful real-world transmission documentation through careful exposure tracking, yet were conducted when ventilation systems differed considerably from modern systems—lacking advanced filtration, air exchange capabilities, and standards now common in healthcare facilities.^22,23^ Inconsistent measurement of environmental factors is evident across the observational studies. While these articles collectively support the persistence of measles in the air between 29 and 120 minutes, their age and methodological limitations suggest additional research is needed to more accurately inform modern contact tracing guidance.

## Discussion

All five hallmark articles that were identified through our search strategy highlight important features of measles airborne infectivity, though limited in scope. Based on the combination of formal and informal search strategies, this search is comprehensive and demonstrates that there is minimal published evidence to guide precise determination of how long measles remains infectious in the air after an infected individual is no longer present. It is clear there is a gap in existing evidence on the duration of measles infectivity in air, and whatever existing research exists is long outdated.

### Study Limitations

Search criteria in the systematic review were limited to manuscripts published in English, potentially excluding useful research in other languages. The search was limited to January 1988 to July 2024 after PCR testing became widespread to reduce potential bias from improper diagnosis, but may have inadvertently excluded earlier studies that could have provided valuable insights into airborne transmission. However, our supplemental search mitigated this risk by including earlier studies. Also, changes in Medical Subject Heading (MeSH) terms over time may have resulted in the omission of relevant studies that were indexed under our search terminologies. Additionally, certain older papers lack abstracts or are available only as scanned PDFs without searchable text, which would not have been detected by the search engines utilized and would have led these papers to be inadvertently excluded. Furthermore, while we developed a comprehensive set of search terms, it is possible that some relevant studies were missed due to variability in terminology used by authors. These factors may have limited the ability to capture the full breadth of all available relevant evidence.

### Research Needs

Health departments currently rely on CDC measles exposure guidelines based on research from De Jong,^8,9^ Remington,^12^ Bloch,^11^ and Sienko^10^ between 1964 and 1987. While beneficial, additional evidence is needed to better understand how long measles remains infectious in the air after a case is no longer present. This evidence can be gathered through additional laboratory and observational studies.

Laboratory studies are needed to better understand how the measles virus survives in the air and on surfaces under different environmental conditions and in various settings. Current literature only evaluates measles survival at 20°C (68°F) and fails to consider the diverse factors present in public settings. Additionally, existing studies have only examined measles survival in controlled environments. Future research should investigate how measles survives in spaces that mimic real-world environments, such as classrooms, open-concept offices, grocery stores, restaurants, airplanes, and hospitals. This will help inform how spatial obstacles and modern HVAC and air filtration systems can influence the movement and survival of the measles virus in places where people are most likely to be exposed. Some modeling studies have addressed this to a limited degree. Riley et al. in 1978 developed a mathematical model to analyze a measles outbreak in a suburban elementary school, highlighting the airborne transmission of the virus.^24^

Additional observational studies gathered from real-world outbreaks are also needed to understand the actual exposure window for measles contacts after cases are no longer present. This information could be gathered through coordinated efforts by state and local health departments conducting measles case investigations in their jurisdictions. Health departments should consider collecting exposure time windows for each contact identified in a case and organizing databases so that epidemiologists can easily determine which contacts became cases. Coordinating entities could then pool de-identified data on the exposure windows for each contact and calculate the attack rates for exposure times.

This systematic review also identified a gap in evidence necessary for contact tracing in outdoor public spaces. Although this systematic review was not limited to indoor settings, the researchers also did not identify any experimental or observational studies that assessed measles virus infectivity in outdoor air spaces that met the other inclusion criteria. Future studies in these settings are needed to inform evidence-based measles contact tracing.

## Public Health Implications

Research is needed to clarify how long the measles virus remains transmissible after an infectious case leaves a public space. Robust evidence would enable health departments to refine and standardize evidence-based guidelines rooted in science and tailored to actual risks to optimize contact tracing while minimizing unnecessary interventions. Whether the research confirms the two-hour window or suggests adjustments, such research would improve public health responses’ efficiency and effectiveness, fostering trust in evidence-based recommendations.

## Data Availability

All data produced in the present study are available upon reasonable request to the authors

## Acknowledgments

The authors acknowledge our state and local health department partners for inspiring this research. We would like to especially thank those health departments that shared the contact tracing guidance they would follow during a measles outbreak, including the Alameda County Health Department, Chester County Health Department, Los Angeles County Department of Public Health, Marathon County Health Department, Public Health-Dayton & Montgomery, Public Health-Seattle & King County, South Carolina Department of Health, Tennessee Department of Health, and additional state and local health departments that remain anonymous. The research team would like to acknowledge the CDC Insight Net Program for supporting this work. The funders had no input or oversight over the conduct of this work.

